# Dissemination of health research results to study participants – a systematic review evaluating current global practice and implications for future research

**DOI:** 10.1101/2025.03.03.25323077

**Authors:** Mary Bagita-Vangana, Holger W Unger, Kamala Thriemer

**Affiliations:** University of Melbourne, Faculty of Medicine, Dentistry & Health Sciences, Melbourne, Australia; Port Moresby General Hospital, Obstetrics & Gynaecology Division, Port Moresby, Papua New Guinea; Global and Tropical Health Division, Menzies School of Health Research and Charles Darwin University, Darwin, Australia; Department of Obstetrics and Gynaecology, Royal Darwin Hospital, Darwin, Darwin, Australia; Department of Clinical Sciences, Liverpool School of Tropical Medicine, Liverpool, UK; Department of Infectious Diseases, University of Melbourne, Doherty Institute, Melbourne, Australia

## Abstract

**Background:** Disseminating research findings to study participants is emerging as a critical component of clinical research. There is evidence that dissemination strengthens social relations and understanding between researchers and participants and their communities, and it is increasingly required by funding bodies. However, there is limited funding support for and guidance on the implementation of dissemination of research results to study participants.

**Methods and Findings:** We conducted a systematic review to describe the current global practice of dissemination of aggregate research results to study participants. The databases Medline (OVID), Embase and CINAHL were searched to identify publications published from January 1, 2008, to March 18, 2024. A total of 87 studies met the predefined inclusion criteria: 37 were qualitative, 29 were quantitative, and 21 were mixed-methods studies. Most studies concentrated on dissemination of broad health-related research (n=25; 26%), followed by cancer research (n=17; 20%) and genetics (n=16; 18%).

Most participants expected researchers to share results regardless of study outcomes. Many participants viewed receiving results as essential for fostering trust with researchers, feeling valued for their contributions, and fulfilling ethical obligations. Many researchers saw sharing results as a moral duty, especially when participants had limited access to scientific knowledge. The most common method for disseminating results was mailing lay summaries or result letters to participants. Group presentations and workshops were predominantly used in lower income countries. Identified barriers and enablers to result dissemination included researcher attitudes and communication skills, logistics and resources, institutional guidance, and ethical and cultural considerations. Impact of dissemination on research findings included improving health literacy, increased understanding of research, and trust in research.

**Conclusions:** Result dissemination is emerging as an integral component of modern clinical research practice and appears to translate into a broad range of benefits in most circumstances. The current lack of agreement on what constitutes best practice will need to be addressed. The design of frameworks to guide the conduct of dissemination, which are now in early development, require validation in a range of settings, populations and clinical domains. Further work on approaches to dissemination of research findings in lower-middle income countries is required.

## Introduction

Disseminating research findings to the research community, which includes study participants, is part of community engagement. Community engagement, a process that involves strengthening social relations and understanding between researchers and participants and their communities (1–3), is recognised as a critical component of medical research (4).

Disseminating research findings is key to research integrity, specifically promoting transparency and discussion of new knowledge, and having the potential to change practice at all levels (5). Indeed, the World Medical Association (WMA) Declaration of Helsinki states that “*all medical research subjects should be given the option of being informed about the <u>general</u> outcome and results of the study*” (6). It further specifies the ethical obligations of researchers, sponsors, and editors to make results available and accessible in the public domain, irrespective of study outcomes (6). Whilst not mandatory, dissemination of research results is now recommended by many research ethics committees and funding agencies, and included in research regulations in Australia, Canada and the United Kingdom (7,8,9). This underlines an emerging imperative that study participants receive aggregate research results.

In 2008, a narrative review summarised the available literature on disseminating research results to study participants (10). It highlighted that research participants want results made available to them, but reported uncertainty about best practise (10). Since then, the number of articles reporting on methods and experiences with results dissemination has increased. Additional work focussing on specific participant populations, geographical locations, fields of research and methodologies are now also available. For example, a systematic review focused on results dissemination in community-based participatory research (11), and a scoping review summarised the experiences with results sharing among Indigenous populations in the Circumpolar North (12). A further scoping review focused on dissemination of study results to participants of phase 3 randomised controlled trials and identified a largely *ad hoc* approach to dissemination by investigators, suggesting a guiding framework is needed (13).

This systematic review aimed to summarise the current global practice of disseminating information on the general outcome and results of clinical research studies (aggregate research findings) to participants involved in health-related research, with specific focus on the following research questions: i) what are the expectations of participants regarding result sharing; ii) what are the reasons for results dissemination; iii) to what extent are results shared with participants; iv) what are the methods used for sharing results; v) what are participant and researcher preferences for how results should be shared; v) what are potential barriers and enablers to disseminating results; vii) what is the best timing for dissemination activities; and viii) what are the impacts of results sharing.

## Methods

### Search strategy and study selection

The databases Medline (OVID), Embase and CINAHL were searched separately to retrieve publications on dissemination of research results to study participants published from January 1, 2008, to March 18, 2024. The search commenced at the cut-off date of a previous review by Shalowitz in 2008 (10). The search strategy included the main keywords ‘results dissemination’, ‘research findings’, ‘study participants’ combined with “OR” and “AND” operators using different keyword variations. The final search string, and the search strategies for each database are presented in Supplementary file 1. Retrieved articles were exported to Covidence (Melbourne, Australia) for screening and data extraction.

### Inclusion and exclusion criteria

Primary quantitative studies including randomised control trials, retrospective or prospective cohort, cross-sectional, case-control or case series studies or studies within a trial or nested cohort studies that reported on reasons, extent, methods of result sharing, preferences by participants and researchers, barriers and enablers, timing of dissemination, and impact were included. Primary qualitative studies were included when they reported on researcher or participant attitudes, knowledge, experiences and perspectives on sharing research results, barriers and enablers, and methods of dissemination. Mixed methods studies that reported any of the predefined outcomes were also included. Inclusion was restricted to articles in English language. Review articles, study protocol, conference presentations and proceedings, guidelines and policy documents, editorials, letters, viewpoints and commentaries were excluded (Supplementary file 2).

### Eligibility of studies

Two reviewers separately screened the articles by title, abstract and full text. Any disagreement between abstract screeners on the eligibility of included papers was resolved through discussion among all three authors.

### Data extraction

The thematic framework for data extraction was informed by prior publications on dissemination and developed prior to extraction. This included location and year of study and whether quantitative or qualitative methods were used. The remaining extractions fields corresponded with the research questions.

At the beginning of the data extraction process, data were extracted by at least two authors (MBV, KT, HWU) to ensure common understanding of the data categories. Data extraction for the remaining articles was then conducted by one reviewer (MBV). Authors of included studies were not contacted for further information or verification.

### Quality assessment and data synthesis

Given the large heterogeneity of study methodologies included in this review, no standard tools to assess quality were deemed suitable. Particularly for qualitative and mixed methodologies the value of critical appraisal is contested (14). We therefore only included study design and sample size as a measure of quality (Supplementary file 3). No studies were excluded based on quality.

### Data synthesis

Results were grouped thematically for descriptive analysis based on *a priory* identified themes. These themes included expectations of participants, reasons for results dissemination, extent of result sharing, methods used, participant and researcher preferences for how results should be shared, barriers and enablers, timing for dissemination activities, and impacts. Quantitative data were not pooled due to significant heterogeneity among study settings, types of research, data collection instruments, and sampling.

## Results

After removing duplicates, a total of 1,138 studies were screened for inclusion. After excluding 919 studies based on title or abstract, full texts were retrieved for 219 studies. A total of 87 studies met the predefined inclusion criteria (Figure 1, Supplemental file 3).

**Figure 1:** PRISMA flow diagram

Of the 87 studies, 37 were qualitative (43%) (3,15–50), 29 were quantitative (33%) (5,51–78), and 21 (24%) were mixed-methods studies (79–99). Most studies concentrated on dissemination of health-related research more broadly (n=25; 26%), followed by studies reporting from cancer research (n=17; 20%), genetics (n=16; 18%), and adult medicine (n=14; 16%). Other research areas included infectious diseases (n=8; 9%), obstetrics (n=4; 4.5%), paediatric/adolescent medicine (n=3; 3%), geriatrics (n=1; 1%) and gynaecology (n=1; 1%). Half of included studies (n=44; 51%) were conducted in the US and/or Canada, and 35 (40%) in the US alone. A total of 25 studies (29%) came from Europe and 8 (9%) from Africa. Three studies (3%) were conducted in the Western Pacific region (Australia, New Zealand and Japan), and four (5%) studies involved multi-national populations.

### What are participants’ expectations?

Thirty-seven studies reported on expectations of study participants about results sharing, a third of them using qualitative methods (n=13, 35.1%), nearly half using quantitative methods (n=18, 48.6%) and 6 studies (16.2%) using mixed methods research. The expectations were described as general expectations (3,15,27,31,35,36,38,42,43,46,48,50,52–56,58,60,62,63,65,66,68,69,72,74,75,78,79,95,98,99), and expectations on aggregate versus individual study results dissemination (30,36,54,58–60,87,90).

#### General expectations

Participants’ views were that researchers should share results with study participants irrespective of study outcomes and response rates ranged from 60% to 100% (15,27,36,48,50,52–56,58,60,62,63,68,72,74,75,79,95,99). The expectation to receive results was associated with a higher level of education among research participants (53,65,78). Participants felt results should be presented neutrally, sensitive results should be shared in an appropriate manner and that results should also be shared even if there was some initial lack of understanding of what the results meant for clinical practice (31,42,69).

#### Returning individual vs aggregated results

Among UK cancer trial participants, 70% (n=57) requested summary results, 69% (n=56) individual results, and 43% (n=35), both. About one quarter requested only overall (n=22, 27%) or individual (n=21, 26%) results (54). Similar findings were found in a Dutch genetics study of 1,678 respondents; 59% of participants were interested in receiving individual results, even if they had no health consequence (90). In a diabetic self-management study, almost all participants (98%; total n=111) desired both individual and aggregate results to influence future health behaviour (36). Whilst there is growing evidence on the strong interest amongst participants to receive individual results, especially in cancer and genetics research, some studies indicate that this will have significant resource implications (30,58–60).

## What are the reasons for disseminating results to study participants?

### Participants’ perspectives

A total of 19 (22%) articles were identified for this theme. Two-thirds of the studies (n=13; 68%) used qualitative research methodologies. Participant reasons for wanting to receive results fell into five main categories: i) to foster trust and transparency (21,30,42,45,66,80), ii) to be recognised for contributions to research (30,31,41,42,45,80,99), iii) as an ethical and moral obligation (3,45,58,59,99), iv) to increase knowledge and awareness (16,22,30,50,58), and v) to improve health decision-making (22,30,32,35,36,42,45,58,66). Two studies reported reasons for not wanting to receive study results (59,60).

#### Fostering trust and transparency

Participants felt that receiving research results would improve relations with researchers and motivate participation in future research (30,42,45). In a cross-sectional survey of 502 study participants in the US, transparency and accountability from researchers were reasons for wanting research results (66), and was identified as a logical conclusion to the research process (21). Respondents in a qualitative US study explained that providing samples and information for research gave them ownership of research findings and therefore sharing final results with them honoured their ‘contract’ with researchers (42).

#### Recognising contributions

Participants also wished to be acknowledged for their time and individual contributions to research (30,99). Being acknowledged made participants feel valued and showed that their participation mattered to the research community (42,45). The act of ‘giving back’ results was also seen as a mark of respect for the community from which participants originated (31,41).

#### Ethical and moral obligation

In a Malawian bioethics study, many participants felt it was their ethical right to receive research results (3). In a survey of adult participants in a cancer study in Canada, 91.5% expressed that they had a ‘strong/very strong’ right to receive information on final study outcomes (58). Similarly, a large survey of parents of children or adolescents with cancer in the US and Canada found that parents felt it was their right to receive research results (59). Community members also viewed receiving results as a moral and ethical obligation for researchers, thereby acknowledging participants’ time and involvement for little or no compensation (45,99).

#### Increased knowledge and awareness

Receiving results for their own education, interest and scientific curiosity were reasons identified in two qualitative studies (22,50). Other participants wished to know future directions of research and how conclusions and recommendations were derived from research data (22). A majority of Canadian cancer research participants indicated receiving results to increase public awareness around cancer research (58), whilst in a US genetics study patients wished to receive results to increase their awareness of scientific progress (30).

#### Improve health decision-making

Returning research results provides information to help personal health decision-making, behaviour modification and health management (22,35,45,66). For example, in genomics studies in Kenya, returning results to the community was perceived as being enabling and motivating for individuals to know their health status and seek care if necessary (30). For cancer patients, respondents in a Canadian study felt the information could help improve their quality of life or reduce risks of harm later in life (58). Similarly, in a qualitative US study of diabetic trial participants, understanding how the research was affecting their community was important to 98% of participants (36). Overall, many study participants recognised an individual as well as collective health benefit (22,32,42) in results sharing.

#### Not wanting to receive study results

Two studies included data on why results should not be shared (59,60). The main reasons cited in a cancer study were distress due to harm as a result of research participation, loss of a family member, and insurance or employment concerns (60). However, many parents of children and adolescents with cancer thought there were no good reasons to not want results (59).

### Researchers’ perspectives

Reasons for sharing results with participants by researchers were described in 14 (16%) articles. Reasons were categorised as follows: i) ethical and moral obligations (3,33,40,45,67,81,88,91,97), ii) recognising participant contributions (45,52,77,81,88,91), iii) fostering transparency and trust (33,45,81,88), iv) improving participant health and research literacy (45,67,91,97), and v) improving individual and community health (37,39,40,88). Four articles described various reasons for not sharing results (33,49,88,97).

#### Ethical and moral obligations

Many researchers considered sharing results a moral and ethical obligation (3,33,40,45,88,91,97). Genetics researchers indicated that failing to communicate results of potential benefit was in violation of ethical principles (40). Others felt an obligation to disseminate results to study participants because they recognised that participants often have limited access to scientific events and journals (88).

#### Recognising participant contributions

Recognising participants’ contributions to research was identified as an important rationale for results sharing (17,88). Acknowledging participants’ contributions to scientific evidence was important even when interventional studies ultimately did not report benefits (45). For example, living elderly participants were informed of the non-benefit of a dietary supplement on cataract prevention after a trial that lasted 13 years (77). Investigators recognised that giving back results acknowledged the important role of participants’ contribution to research and although disappointed, participants felt that their contribution mattered enough for investigators to return results to them (77).

#### Fostering transparency and trust

In a mixed-methods study amongst US researchers, returning research results was found to build relationships and trust between researchers and participants (88). Returning aggregate results was a way of reciprocating participants’ engagement with research (88) and accounting for public funds used for research (33,88,91). The dialogue and exchange of information created a sense of shared ownership and value among participants which encouraged research participation and new ideas for research (33,45,88).

#### Improving health and research literacy

In a survey of genetic researchers, 87% of respondents felt it their duty to ensure participants were aware of the latest developments (67), whilst in HIV and malaria research in LMICs, returning results helped researchers to correct errors, misinformation and myths about infectious diseases of public health importance (91,97). Furthermore, returning results was thought not only to satisfy scientific curiosity but also to improve health and research knowledge of individuals and communities (97).

#### Improvement of individual and community health

In a community engagement study in Brazil, returning results to participants and their communities appeared to strengthen HIV-related care processes, thereby overcoming some challenges participants experienced in a fragile healthcare system (37).

#### Reasons for not sharing results

Some researchers argued that aggregate study results should not be shared with participants (33,49,88,97). For example, 15.7% of US health researchers surveyed disagreed with dissemination and a further 19.8% were unsure (88). Principal reasons given were that the health literacy of participants could limit their understanding of research findings; participants did not explicitly ask for results; there were challenges with explaining inconclusive/incomplete results; and concerns about potential harms to participants may be triggered by sharing results (33,88). The reason that participants did not always ask for results appears to underscore the power inequality between researcher and participant as it assumes that participants may not be interested (97). Other reasons to not disseminate aggregate results included concerns that this could cause harm to research; foster misconceptions about clinical trials which may ultimately not always report benefits for participants; and violation of privacy in small communities where participants may be easily identifiable (33,49,88). Some researchers thought publication in peer-reviewed journals was sufficient and where participant involvement was minimal, there was no need to disseminate results (88).

### To what extent are results disseminated to study participants?

A total of 45 articles focused on extent of results dissemination to participants. There were slightly more quantitative (n=18, 38%) than qualitative (n=16, 34%) and mixed methods (n=13, 28%) studies. Of these studies, 32 (68%) disseminated results directly to participants - 26 (55%) from a parent trial (16,17,21,23,24,34,36,39,41,46,48,52,55–57,60,64,65,69,77,80,82,85,86,91,95) and 6 (13%) from general surveys of researchers and participants (28,62,63,79,81,88). Thirteen (32%) studies were not related to direct dissemination of study results to participants but included results relevant to the theme (5,18,30,33,35,54,58,70,71,83,89,94,97).

Many health researchers considered but did not widely practise disseminating results to participants after study completion (35,58,89), the process itself being described as suboptimal by one author (83). In a survey of 414 US researchers, two thirds thought research results should always be shared; however, only 8.1% had specific plans for sharing (88). Among malaria researchers, 83% thought results sharing was ‘important, but only 25% conducted such activities after trial conclusion (97). In another survey of 79 trial participants, the majority (86%) had not been asked if they wanted to receive research results (54). Further evidence of the inertia to share results with participants was seen in a large UK audit of phase 3 registered clinical trials in which 12% (n=173) of 1,404 registered trials did not have explicit plans to share results with participants. While 88% (n=1,231) did intend to disseminate results to participants, only 19% (231/1,231) actively planned sharing results and 81% depended on passive means requiring participants to seek aggregate study results on their own initiative (71). These and other examples show a clear gap between intention and behaviour regarding result sharing (30,33,35).

A systematic review investigating community-based participatory research (CBPR) in the US in 2010, found that contrary to poor dissemination to participants in clinical trials (71), dissemination to participants and the general public in CBPR was largely occurring, with 98% of 63 authors reporting dissemination to community participants and 84% to the general public (11). However, among National Health Institute (NHI) - funded studies in the US, less than 20% of researchers shared results with community representatives (28).

In a review of 238 patient information leaflets (PILs) in Europe (18), 74% described plans to share results with participants; however, the majority relied on participants’ own initiative to find results. In addition, 90% of the planned dissemination exercises were largely suitable for health professionals rather than participants (18). Another European study of plain language summaries (PLSs) found that only 14 of 99 (14%) trials had PLSs available in the public domain and that these were not easy to find on internet search engines (70). In a survey of 1,818 PubMed authors, 27% (n=498) shared results with participants, with half using lay summaries sent to patient groups/communities, patient-friendly conferences, mainstream media or online (5). Furthermore, an assessment of 48 trial protocols in LMICs identified that 44% contained communication language but none included communication plans (94).

### What are the methods used for disseminating results?

Fifty studies described methods used to disseminate research results to participants. Most (n=20, 40%) were qualitative studies, followed by 36% (n=18) quantitative and 24% (n=12) mixed-methods studies.

The most common format reported was the posting of lay summaries or result letters to participants (52,55,56,65,69,71,72,77,80,85,86,95). To illustrate, in a British randomised controlled trial among survivors of acute myocardial infarction, 80% of those who requested results received lay summaries via post (55) and in a breast cancer trial in the US, 304 Spanish-speaking Latino participants received summary results via standard mail (52). Other studies that utilised mail to send lay summaries, leaflets or results letters were to parents of children with advanced cancer (41,80), women in a prenatal antibiotic trial (16,48,85), and elderly participants in a cataract prevention trial (86). Some of the posted summaries included a telephone number for questions (56,86) and a website link (86).

Another common mode of dissemination was group presentations. This was done in a number of ways, including PowerPoint presentations for men involved in urological studies (17,60), roundtable meetings with community members in an Ugandan village (33) and among Hmong participants (27), dissemination workshops in Malawi, Brazil, and Kenya (3,37,41,70), using pictorial aids in local languages for Ethiopian malaria trial participants (24) and paediatric participants and their parents (76), and classroom discussions among year 11 students in South Africa (91). A photo essay workshop for Huntington disease participants and their families was organised to engage with them on sensitive issues (19).

Other modes utilised were email notifications (23,61,63,79,81), websites (44,51,64,82), telephone calls (57), and individual face-to-face meetings (21,34,36,79,83,90). Several studies reported multiple strategies to address various needs of community members (5,18,35,39,45,58,97,99). Other creative methods employed were a comic (61) and a short film (47).

## What are the preferences for how dissemination of results is conducted?

### Participants’ perspectives

Thirty-four articles evaluated participants’ preferences for dissemination. The majority were quantitative (n=15, 44%) followed by qualitative (n=12, 35%) and mixed-methods studies (n=7, 21%). A third of these studies (n=11, 32%) focused on dissemination preferences among cancer patients.

The preferred format in many reports was text media including, but not limited to, lay summaries, leaflets, results letters, brochures, and posters (15,22,33,42,50,52,55,56,58,59,65,66,66,68,72,74,75,79,80,86,90,98,99). For example, in a cardiac rehabilitation study, 80% of participants wished to receive results by letter (55) and among 32 adolescent participants with idiopathic scoliosis, 95% wished to receive results in the written form either by letter or a brochure or pamphlet (56). Many participants preferred to receive these text media via standard mail. Among French participants in breast cancer genetic studies (65) and older participants in a qualitative preferences survey (42), postal delivery allowed participants to have a ‘hard copy’ on hand to refer to or share (79,98).

Other studies reported preference for electronic versions of lay summaries. For example, receiving online summaries via email was preferred by cancer participants in a US survey (79) and 65% of French patients with a rare disease (15). However, participants in some studies did not favour websites to access online material (34,44,90).

Fewer publications indicated group meetings, workshops or other interactive methods as preferences for receiving study results (30,35,41,44,54,60,98).

Less commonly reported formats were dissemination of study results through mainstream media (33,56,90) and pictorial representation in the written or electronic form (27,52). Preference for a comic over lay summaries and scientific abstracts was described in a randomised controlled trial (61), and a film was preferred for communicating HIV research to young people (47).

Other formats reported were via telephone calls (15,63), meeting with other parents (15), digital video disk (DVD) (98), twitter and text messages (63), and scientific publications (42,61).

Many participants preferred a combination of formats and modes to receive results, for example, a telephone number in their results letter (56,99), a link to an online webpage in their letters or emails (56), a meeting after receipt of results (22), or dissemination tailored to the needs of the study population (83).

### Researchers’ perspectives

Nine articles described researcher preferences for results dissemination to participants. Of these, 22% (n=2) were quantitative studies, 44% (n=4) were qualitative and 33% (n=3) were mixed methods studies.

A survey among research investigators indicated that 44% preferred lay summaries followed by info-graphs (39%) and newsletters (39%) to share results of their studies (45). Other preferred methods were via online platforms (45,71) and group presentations (17). While the use of social media was a potential avenue for malaria researchers (97), others preferred to use multiple strategies including social media, lay summaries, a letter with a number to call, and online platforms depending on context (35,45,54,97).

Tailoring communication plans to participant preferences and population characteristics, for example, employing print material and standard mail for older participants and online and social media for younger participants, were reported (45). Five studies reported that researchers considered community and participant engagement important in planning for dissemination (35,40,45,91,99).

## What are the barriers and enablers to dissemination?

### Participants’ perspectives

Twenty articles described participant barriers and enablers to dissemination of study results. Sixteen (80%) were qualitative studies, 3 (15%) were quantitative studies and 1 (5%) a mixed-methods study.

Barriers and enablers suggested by participants were grouped as: i) researcher attitudes and communication skills (20,22,25,29,35,38,54,59), ii) logistics and resources (22,34,35,38,42,43,53,54,59), iii) personal circumstances (17,59) and iv) participant engagement (3,25,30,35,41,42,45,47,50,53,99).

#### Researcher attitudes and communication skills

A condescending attitude was perceived as a significant barrier by participants in patient-centred outcomes research (22), often leading to ineffective communication (20,29,38) and poor understanding of information by participants (59). This included the non-prioritisation of communication with participants by researchers and a lack of awareness and insensitivity to participants’ needs (22,38). At the same time, staff being able to understand context was considered enabling, for example, understanding why participants in cancer research want feedback (54) or understanding cultural contexts and being culturally appropriate toward Indigenous Alaskans (25) and Afro-American and Latina participants (35).

#### Logistics and resources

Researchers’ time constraints (22), participants’ lack of time (59), and the burden of committing to a trial (38) were noted as barriers to dissemination. Cost was of less concern to Canadian parents of paediatric cancer patients (59). Providing extra funding for research and staff (42) to cope with the workload, and appropriate training for research staff, were identified as enabling factors among African American and cancer participants (34,53).

#### Personal circumstances

Emotional challenges grounded in personal circumstances were identified as barriers to dissemination in paediatric cancer research. The death of a child would make it more difficult to desire results, as it evoked memories of the illness (59).

#### Participant engagement

Participants criticised a lack of community engagement by researchers and suggested involving participants in the dissemination process (30,45,50,53,99). In Malawi, study participants complained that only village chiefs were consulted before study commencement (3). Participants in a Kenyan HIV study dissemination workshop desired consideration as active partners. This was reflected by suggestions to train community members in relaying research findings to the wider community, thus helping to change behaviours and reduce spread of the infection (41). Similarly, collaborating with trusted community leaders would enable researchers to disseminate study findings in appropriate ways (42). Co-development of a film and leaflet by teenage participants with perinatal HIV for other affected young people (47) and collaborative efforts with indigenous and ethnic minorities enabled more effective dissemination of results (25,35).

### Researchers’ perspectives

Thirty-one studies described researcher barriers and enablers to research dissemination. Half the studies were qualitative (n=16; 52%), one third were mixed-methods studies (n=10; 32%) and the rest were quantitative studies (n=5; 16%). Barriers and enablers for researchers included i) reaching/contacting participants (3,19,33,35,40,77,81,97), ii) limited preparedness (35,45,88,89,97), iii) logistics and finances (17,24,33,35,39,43,50,54,60,72,81,86,88,89,91,94,96), iv) continuous communication (5,30,33,35,40,41,44,45,54,81,83,89), v) institutional guidance (5,33,35,40,41,81,88,89,94,97), and vi) ethical and cultural considerations (5,37,39,45,49,88,89,93).

#### Difficulty reaching participants

In a survey of US and Canadian genetic counsellors, 31% of respondents reported that after trial involvement was complete, it was difficult contacting participants (81). Similarly, research staff involved in bioethics research in Malawi noted that many participants were not contactable and may have relocated during the long interval between end of study and availability of results (3). In Italy, older participants often appeared to change addresses and phone numbers (77). Many researchers reported low literacy levels among participants in LMICs as being a barrier to reach participants. This was perceived as contributing to inabilities to comprehend research results (40,97) even if translated to local languages (40). Time constraints, lack of interest, mistrust, and being too academic also made dissemination more difficult (19,33,35,97).

#### Limited preparedness

Lack of early planning at study inception, together with long intervals between trial completion and completion of the final report, were reported as the main barriers among malaria researchers (97). As a result, not planning ahead could lead to poor or no dissemination or an expensive exercise (45). In addition, lack of awareness about returning results was reported as the main barrier among US health researchers (88,89). Additionally, researchers often felt they lacked time or communication skills (35,89) to effectively impart results to a lay audience.

#### Logistics and finances

Logistic barriers were reported by a majority (68.9%) of health researchers in one study (88). These included inability to contact participants as contact details were either not collected, or rural/remote location precluded internet or phone contact (39,88,89). Lack of electricity and poor road conditions were additional challenges in Uganda (33). Furthermore, the lack of communication methods that are cost-effective and time-efficient (86) such as challenges with organising group events (24) or time spent disseminating printed materials were identified as barriers (43,72,96). The lack of specific funding for dissemination activities was identified as a major obstacle (35,50,88,91). Among malaria researchers in LMICs, 52% identified inadequate funding as a barrier to dissemination (94) and among researchers in the US, 54% reported financial constraints that had impacted on their ability to disseminate findings to participants in previous studies (89). Allocating extra resources, such as data-tracking systems to allow long-term follow-up, were reported as facilitators by 12.8% of cancer researchers (54). Similarly, finding an inexpensive but appropriate venue and keeping group presentations simple worked for a Canadian dissemination exercise (60).

#### Continuous communication

The initial consent process was thought crucial by researchers to discuss the concept of dissemination to participants. For example, in a survey of UK cancer researchers, 32% of respondents indicated the importance of early communication about end of trial result dissemination to better understand participants’ preferences (54). Furthermore, 18% of participants in this same study suggested that transforming dissemination activities into a routine task would improve dissemination (54). Researchers thought community members play an important role in dissemination and should be engaged effectively as partners in developing and implementing dissemination plans (33,35,40,41,83). Understanding terminology surrounding return of results has also been identified as important. For example, Kisiangani suggested using terminology like ‘sharing knowledge’ rather than ‘returning results’ when referring to the return of aggregate results (30). In a French qualitative study of breast cancer patients, the term ‘results’ was generally misunderstood with difficulties distinguishing between results related to medical research and those of usual clinical care (44). Differentiating between ‘aggregate results return’ and ‘dissemination of findings’ was suggested (45). Overall, agreement on definition of terms would enhance participant and researcher understanding.

#### Institutional guidance

Some researchers reported a lack of guidance at national and institutional level as an additional barrier (5,35,40,41,81,89,94,97). Only 22.5% of malaria researchers received institutional support and 8% thought institutions lacked interest in disseminating to participants (97). Moreover, there was a lack of dissemination requirement from funding bodies to the extent that sharing results with participants was perhaps not seen as an essential component of the research process and funding (89). Other institutional barriers included lack of communication and training (35,40) and lack of incentives (5,33,35,88).

#### Ethical and cultural considerations

Ethical concerns were reported as barriers among 38.5% of US studies (93). Three main concerns were that of harm caused by either emotional distress or stigma arising from learning about research results (93), misunderstanding or misinterpretation of results (5,88,93), and concerns about maintaining privacy and confidentiality of participants (49,93). Concerns about patient confidentiality were highlighted in Burkina Faso where hierarchical structures and power differences existed within small research communities (49). However, understanding cultural context, using vernacular knowledge and critical pedagogy were strong enablers for dissemination of HIV-related activities in a poor Brazilian community (37). Regulatory barriers included the perception that sharing results required approval by institutional review boards (5,45,88,89).

## What is the best timing of dissemination to participants?

### Participants preferences for timing of dissemination

Eighteen articles described participant preferences for timing of dissemination of research findings. Eight (44%) of these studies were quantitative, 8 (44%) were qualitative and 2 (11%) were mixed methods studies. Participant preferences for timing of results dissemination commonly described the following time periods: i) during the course of the study (3,22,30,41,42,44,51,54,64,75,86), ii) at the end of study before publication of results, with or without peer review (3,22,35,41,42,51,54,57–59,63,75,83), iii) upon publication (51,59,75), and iv) after publication (59,63).

Timing played a key role in understanding participant’s experiences of results dissemination (100). Studies that described participant preferences for receiving results during the course of the study placed emphasis on long-term studies (3,22). The disclosure of results many years after the end of an obstetric study (UK) brought unpleasant feelings of guilt and uncertainty to those who perceived results as being ‘bad’ (85). In other studies, participants involved in cancer and genetics research felt that the time interval to learning results was too long (30,64) and others participating in diabetes and urology studies felt that regular updates helped them overcome feelings of disappointment at receiving results later than expected (86).

Almost three quarters of studies (n=13, 72%) described participants preferred timing for receiving results to be at the end of the study and before publication, either before or after peer review. Preferences ranged from 24% of parents (n=98) and 33% (n=28) of adolescents with cancer (59) to 78% (n=124) of genetics research participants (51). While 64% of 189 adult oncology patients preferred results disclosure prior to peer review, 27% of parents and 15% of adolescents with cancer preferred receiving results after peer review (58,59). In general, participants preferred to be the first, and not the last, to know of study results (51,59).

Preference for receiving results as soon as the study was accepted for publication ranged from 26% of adolescents and 33% of parents of children with cancer (59) to 44% (n=70) of genetics research participants (51). Post-publication dissemination of results to participants was preferred by 12.8% (n=11) of adolescents with cancer and 45.6% of past research participants in two studies (59,63). Other timing preferences described were receiving results soon after preventive measures were available, upon contacting the research team, or within a year of study completion (30,51).

Overall, participants expectations on when to receive results are not well documented. Representing 178 UK trials in Ireland and the UK, only 8% of 238 PILs included a time period for when to expect results while the majority of PILs did not indicate a time period (47%) or did not include any information (45%) on when to expect results (18).

### Researchers’ preferences for timing of dissemination

Studies describing researcher preferences for timing of disseminating study findings to participants were limited. Three studies described preferred timing by researchers for sharing results with participants (3,5,54). In a Malawian dissemination workshop, research staff felt participants should receive results after study completion (3). Meanwhile, in a survey of UK cancer researchers, 89.4% (n=127) endorsed sharing results after a trial had ended and at the time of publication (54). In comparison, among clinical trial researchers across 71 countries, two-fifths had either disseminated or planned to disseminate study findings within 2 years after publication (5).

### What are the impacts of dissemination?

Fifty-five studies described impacts of dissemination on study participants. Almost half the studies were qualitative (n=26, 47%), and a quarter each were quantitative (n=14, 25%) and mixed methods (n=15, 27%) studies. The impacts of dissemination described included: i) evoking emotional responses (16,17,24,27,30,33,35,37,38,41–43,45,48,52,56,59,60,69,72,77,80,82,85,87,95,96), ii) improving health literacy and decision-making (5,23,30,34–37,47,76,86,90,91), iii) understanding research in which they participated (17,56,58,60,77,79,84–86,97), iv) likely participation in future studies (5,15,17,35,36,48,56,84,92), v) trust in medical research (5,41,48,49,64,65,84,89), and vi) impact on the broader community (5,39–41,60,94,99). A lack of post-study dissemination also impacts participants (3,20,21,29,35,45,49,52,66,73,84).

#### Evoking emotional responses

Receiving results evoked mostly positive but also negative emotions. Most common feelings among participants who received aggregated study results were those of satisfaction (42,56,60,69,72,77,80), gratefulness (17,80), and appreciation (24,41). Other positive emotions included being relieved (56), pleased (33,69,95), feeling valued (41,45), respected (35,41) recognised (80) and accomplished (43,45) as well as knowing their contribution was important and valuable (37,43,45,52). As an example, 71% of parents of children with advanced cancer felt ‘a lot or extremely satisfied’ about receiving results (80). In a Canadian study of men with prostate cancer, all respondents were satisfied with the manner in which they received results (60) and 60% of elderly participants in a cataract trial were satisfied receiving results (77). While most participants responded favourably, a smaller proportion of participants experienced negative emotions. These included feelings of regret (48,95) and guilt (56,72,85), anxiety (27,56,69,72,80,85), confusion (38,80,87), sadness (56,72,80) and anger (56). For some women involved in a prenatal antibiotic trial, regret and guilt were strongly felt after participating in a trial that resulted in poorer outcomes for their babies (48). In an ovarian cancer trial, 16% of participants found the results upsetting and 3% regretted learning about the results (95). These findings indicate that while most participants were satisfied receiving results, some experienced negative emotions, especially those receiving unexpected or unwelcome findings, mostly in cancer research. Help and support for participants may be required in these instances (16,30,37,96).

#### Improving health literacy and decision-making

Participants in a diabetes trial indicated that their general knowledge about diabetes improved (23). Similarly, patients in a HIV study also saw an improvement in their knowledge about the infection (37). Others felt that receiving results improved their understanding of their specific medical condition and motivated them to make better health decisions for themselves (30,35,36,76), or for their children (76).

#### Understanding research better

In a range of studies respondents indicated that results dissemination improved their understanding of research when results received were clear, concise and easily understood (56,79,85) and dissemination was described as being informative (56,60,79,97). In a qualitative study, comprehension of research results was better after a presentation compared to reading the scientific publication of an urology trial (17). Among African American participants, receiving results increased overall understanding of the research process (84).

#### Participation in future research

Result dissemination can impact on willingness to participate in future research. For example, 40% of parents of children with a rare disease were willing to participate in future trials (15). Similarly, participants in adult medicine (17,36), minority ethnic groups (35,84), and adolescent health (56) expressed higher willingness to participate after receiving research results. However, some participants expressed they would never participate in a trial again and this was mainly attributed to what they perceived as a negative experience (48).

#### Increase trust in medical research

Among African American participants, trust in researchers significantly improved (84), and 28% of participants involved in breast cancer genetic studies reported increased trust in medical research after dissemination of results (65). Similarly a lack of confidentiality and poor consent processes undermined trust in medical research in an obstetric trial (48,49). In a breast cancer trial, less than half (47%) of participants were able to access research summaries via a website link and while this was intended to increase trust in research, the opposite was achieved (64).

#### Impact on the broader community

Finally, disseminating results to study participants can impact the wider non-academic community. For example, key findings from the dissemination process in an HIV study were adopted to other health research in Kenya (41). In addition, 91% of Canadian prostate cancer participants (60) and those in an Irish hypothyroidism trial agreed that the results would be of benefit to the broader community (99).

## Discussion

Despite increasing awareness of the need and potential benefits of returning research results to participants, further progress with embedding result dissemination into clinical research is needed. Whilst many researchers intended to share results and most participants desired learning about research results, multiple barriers to dissemination persist. Poor planning, funding constraints, lack of institutional support and lack of guidance on best practise were found to be key challenges. Unless these barriers are addressed, efforts to close the dissemination gap will remain challenging. There is a need for researchers to better engage with the communities they work in and tailor communication strategies to participant preferences through co-design. While processes to avoid unethical research practices are well established in relation to consenting and study conduct, there is less guidance or monitoring of post-trial processes including result dissemination. Frameworks to guide the planning of results dissemination are limited (101).

Dissemination practices are reported mostly from middle-to-high income countries in the Americas and Europe. This reflects findings from previous reviews (10–13) suggesting limited progress in relation to the dissemination of aggregate research results in lower-resource settings, where a significant amount of clinical research is now being conducted (102). While there has been an increase in dissemination studies from LMICs, these have mainly come from the African continent. This missed opportunity of engaging with communities to enable high-quality research centred on community needs must be mitigated through the development of guidelines, funding support, and training.

Furthermore, dissemination is currently mostly performed in cancer and genetics research but seems to translate into positive benefits for both participants and researchers across other medical domains. Co-design of clinical research that integrates dissemination of results also appears to substantially improve community acceptance in underserviced patient populations, for whom access to research is limited because of language, systemic racism, gender and poverty (103). Furthermore, dissemination may be a key element to restore trust in medical research and is critical to overcome imbalances in health outcomes.

Participant and researcher preferences for methods of dissemination may not always align, and multiple modalities of dissemination may be required. Dissonance largely appears to arise from financial and logistical constraints to enable researchers to match participant expectations. Furthermore, preferences for how participants wish to learn about study results are invariably diverse as they depend on socio-cultural, economic and disease context. Failure to determine these context-specific requirements at the outset may therefore undermine the impact of any subsequent dissemination efforts. Furthermore, enabling better tracking of participants to permit researchers to reach former study participants in a safe and appropriate manner is essential. This may be facilitated by early planning and funding dissemination of interim results or study progress that keeps participants engaged, including canvassing feedback from participants on their study experience to date.

The present review focussed on summarising the current state of activities relating to the dissemination of aggregate study results. However, our review indicates that study participants may also be interested in receiving individual test results arising from their participation in, e.g., a clinical trial. While at least some individual results generated during trials might be shared with patients in the context of their clinical care, findings from downstream sample analyses are often not shared. It remains to be explored how much individual result sharing is feasible within the context of trials and how much it can potentially foster future engagement in research. Most of the literature on individual result sharing derives from genomic research where interpretation of clinical significance of results remains challenging and complicates the process compared to standard clinical data (104).

Our literature review has several limitations. Firstly, while there is an increase in literature on result sharing, its synthesis is challenging given the substantial heterogeneity in study types, methodology, and outcomes that were assessed in relation to results dissemination. Secondly, reporting bias is likely to influence this review towards more reports of successful or generally positive experiences with result sharing. Lastly, our review was limited to English language publications and did not consider grey literature, potentially missing relevant reports.

## Conclusion

Result dissemination is emerging as an integral component of modern clinical research practice and appears to translate into a broad range of benefits in most circumstances. The current lack of agreement on what constitutes best practice will need to be overcome through the design of frameworks to guide the conduct of dissemination, which are now in early development and require validation in a range of settings, populations and clinical domains. Early planning and co-design appear to lead to more effective dissemination and positive returns as they are more likely to meet community preferences but require adequate funding support. Further work on approaches to dissemination of research findings in lower-middle income countries is required. Lastly, defining outcomes, such as measures of participant satisfaction, engagement in clinical research, and translation of research findings into personal health practices will enhance assessment of the overall impact of dissemination itself, and clinical research overall.

## Author contributions

KT and MB conceptualised the review. MB, KT, and HWU reviewed, extracted and synthesised the data. MB wrote the first draft of the manuscript. MB, KT, and HWU edited and finalised the manuscript.

## Other information

This review was not registered. Protocol information and relevant data is available in the supplementary files. No specific funding was obtained for this work. The authors declare no competing interest.

## Supplementary material

**Supplementary file 1:** Search strategy

**Supplementary file 2:** Eligibility criteria

**Supplementary file 3:** Characteristics of included studies

## Data Availability

This is a review.

