## supplement file 1 for "Dissemination of health research results to study participants – a systematic review evaluating current global practice and implications for future research"

**Supplementary file 1: Search strategy**

**Key words and search string**

The search strategy included the main keywords ‘results dissemination’, ‘research findings’, ‘study participants’ and combined with “OR” and “AND” operators using different keyword variations. The final keyword chain used was: “(result* adj2 (disseminat* or disclos* or feedback or return* or communicat* or inform* or provid* or shar* or reeiv* or send*)) AND (research or trial or study) adj1 (result* or finding*) AND ((trial or study or research) adj2 (participant* or patient* or subject* or volunteer*) OR research community).

The keywords were adjusted depending on the database searched.

a) **Search strategy in Medline (OVID)**

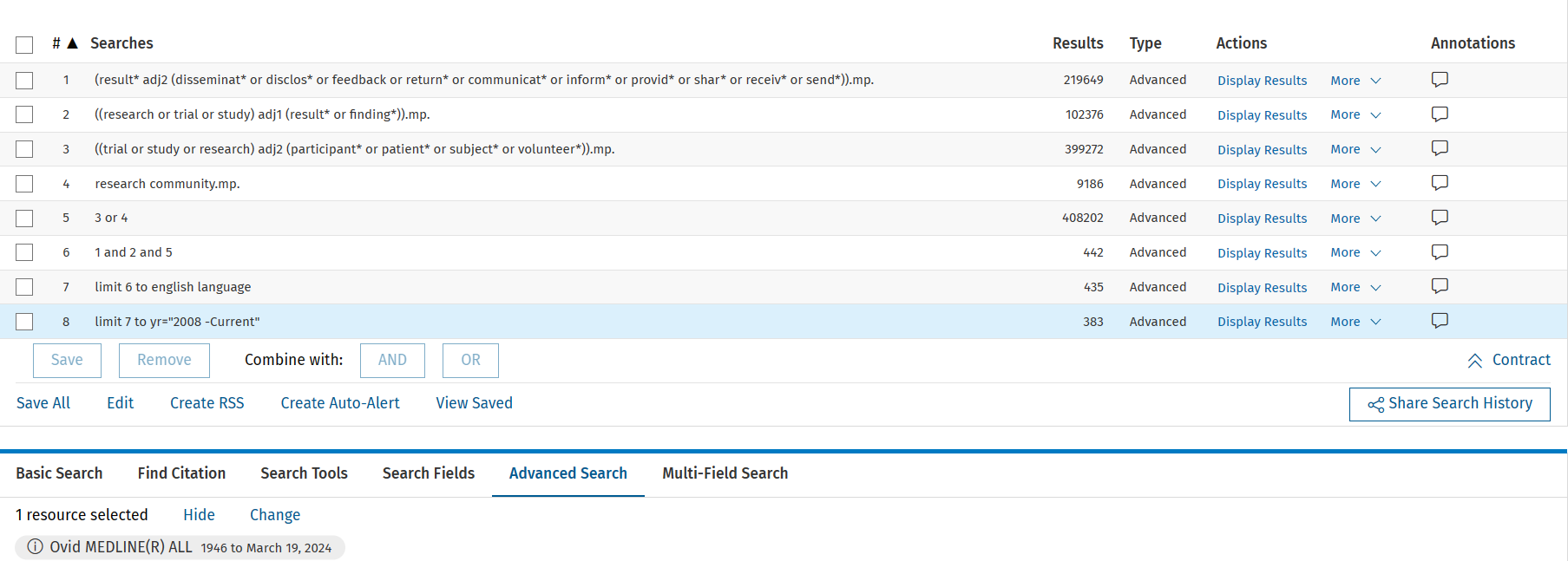

b) **Search strategy in Embase**

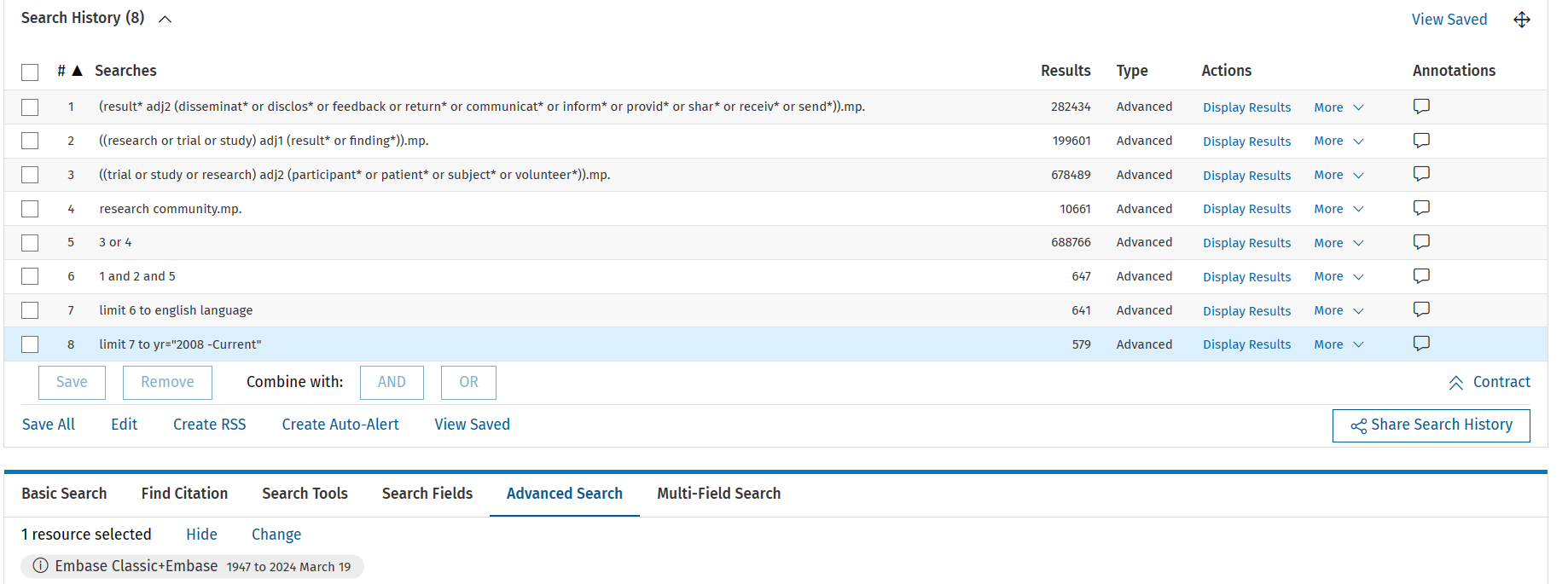

c) **search strategy in CINAHL**

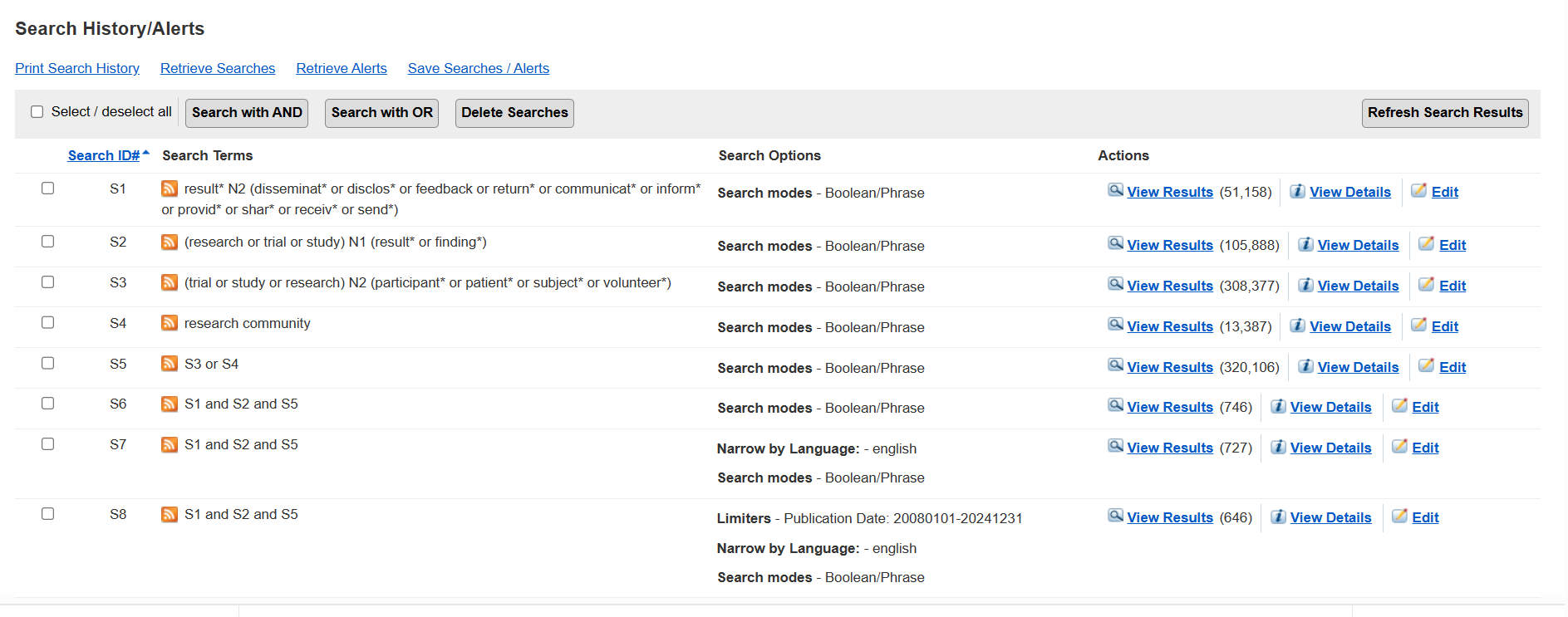
