## supplement file 2 for "Dissemination of health research results to study participants – a systematic review evaluating current global practice and implications for future research"

**Supplementary file 2: Eligibility criteria**

**Inclusion:**

1. Study design - all primary quantitative, qualitative and mixed methods studies:
   1.1 Primary quantitative studies include randomised control trials, retrospective or prospective cohort, cross-sectional, case-control or case series studies OR studies within a trial or nested cohort studies that report the following:
       - Reasons for sharing research results;
       - Extent of result sharing with participants;
       - Modes of research results dissemination to researchers and participants;
       - Participant and researcher preferences for sharing results;
       - Barriers and enablers to communicating research findings;
       - Participant and researcher perceptions on results sharing;
       - timing of dissemination
       - Impacts of result sharing on participants and communities;
   1.2 Primary qualitative studies include those that report the following:

- Researcher or participant attitudes, knowledge, experiences and perspectives on sharing research results;
- Barriers and enablers to sharing results;
- Modes of result sharing.
  1.3 Mixed-methods studies include those that report on any of the above.

2. Exposure of interest - communicating research results to study participants.
3. Reported outcomes - impact on participants and the research enterprise such as positive or negative experiences, knowledge transfer, satisfaction, influence on future trials and research including participant recruitment and retention.
4. Population - all groups: paediatric, adolescent and adult populations including pregnant women and minority groups;
5. Geographical location - global.
6. Setting - any health care facility including hospitals, community health centres, general or nurse practitioner clinics, or within the community setting.
7. Date – January 1, 2008 to March 18, 2024
8. Language - English language only due to time factor.
9. All peer reviewed articles.

**Exclusion:**

1. Review articles (literature, scoping and systematic reviews).
2. Study protocols.
3. Conference presentations.
4. Guidelines and policy documents.
5. Editorials, letters, viewpoints, or commentaries.
6. Non-English studies or those without an English translation.
